# Should we Mitigate or Suppress the next Pandemic? Time-Horizons and Costs shape optimal Social Distancing Strategies

**DOI:** 10.1101/2021.11.14.21266322

**Authors:** Sarah Nowak, Pedro Nascimento de Lima, Raffaele Vardavas

**Affiliations:** University of Vermont, Burlington, VT; RAND Corporation, Santa Monica, CA; Argonne National Laboratory, Lemont, IL

## Abstract

The COVID-19 pandemic has called for swift action from local governments, which have instated Nonpharmaceutical Interventions (NPIs) to curb the spread of SARS-Cov-2. The quick and decisive decision to save lives through blunt instruments has raised questions about the conditions under which decision-makers should employ mitigation or suppression strategies to tackle the COVID-19 pandemic. More broadly, there are still debates over which set of strategies should be adopted to control different pandemics, and the lessons learned for SARS-Cov-2 may not apply to a new pathogen. While curbing SARS-Cov-2 required blunt instruments, it is unclear whether a less-transmissible and less-deadly emerging pathogen would justify the same response. This paper illuminates this question using a parsimonious transmission model by formulating the social distancing lives vs. livelihoods dilemma as a boundary value problem. In this setup, society balances the costs and benefits of social distancing contingent on the costs of reducing transmission relative to the burden imposed by the disease. To the best of our knowledge, our approach is distinct in the sense that strategies emerge from the problem structure rather than being imposed *a priori*. We find that the relative time-horizon of the pandemic (i.e., the time it takes to develop effective vaccines and treatments) and the relative cost of social distancing influence the choice of the optimal policy. Unsurprisingly, we find that the appropriate policy response depends on these two factors. We discuss the conditions under which each policy archetype (suppression vs. mitigation) appears to be the most appropriate.

## 1 Introduction

The COVID-19 pandemic has called for swift action from public health officials, who have instated Nonpharmaceutical Interventions (NPIs) to curb the spread of SARS-Cov-2. Because blunt NPIs (i.e., lockdowns) have far-reaching consequences, the question of how society should use the available instruments to curb SARS-Cov-2 transmission has been met with a deluge of research and controversy.

Broadly, two classes of strategies emerged from this debate: suppression and mitigation strategies. *Suppression strategies* -the strategy adopted in many countries - seeks to reduce transmission as much as possible while treatments and/or vaccines are not available through a variety of instruments, including lockdowns, social distancing mandates, and mask-wearing. Countries known for their strong suppression strategies include New Zealand, Australia, South Asian countries, and others. A *mitigation* (a.k.a. herd immunity, also portrayed as “focused protection”) approach attempts to limit the impacts of the disease on the most vulnerable members of the population while allowing the disease to spread, hoping to balance the costs and benefits of social distancing measures [1]. The argument underlying this strategy is that social distancing would be too expensive to contain transmission relative to the cost of infection for the average person. For COVID-19, suppression strategies have deserved and received overwhelming support in the scientific community due to the high death rates caused by the disease, as well as due to the imminent threat to the stability of health systems and continued uncertainty about the duration of immunity acquired through infection [2]. As the COVID-19 pandemic demonstrated, countries that relaxed suppression strategies too soon (e.g., India, Brazil) did go through worst-case scenarios where health care was severely impacted, and excess death soared [3, 4].

The importance of social distancing policies has motivated an emerging and growing stream of papers evaluating the “lives vs. livelihoods” tradeoffs imposed by pandemics [5–8]. Although empirical work has demonstrated that the short-term economic impacts of NPIs can be limited [9], model-based research evaluating the health and economic effects of NPIs have shown tradeoffs between economic and health outcomes [5, 10–13], and have demonstrated that testing and quarantining can ease these tradeoffs [14]. Many of these articles consider the early stages of a pandemic when a vaccine is not available, and social distancing measures are used to control the spread of the disease. In this paper, we contribute to this literature by demonstrating the role of varying time horizons and perceived costs of social distancing on the optimal social distancing policy.

To better understand lives vs. livelihoods tradeoffs and how it shapes policy and the course of a pandemic, it is useful to combine behavioral models of society with disease transmission models describing the dynamics of how the virus spreads [15, 16]. In this paper, we investigate how the choice of the strategy to govern Nonpharmaceutical interventions change depending on society’s assessment of the time-horizon of the pandemic (i.e., the time it takes to obtain and distribute an effective vaccine) and the costs of suppression measures relative to the social cost of infections. We developed a model that considers the tradeoff between COVID-19 suppression and mitigation strategies based on case counts and social mixing over a pre-specified time horizon.

This paper employs a deductive reasoning approach to modeling behavior in an infectious disease model, where behavior is defined as the solution to a boundary value optimization problem (BVP). This analytical structure (a deterministic SIR model) and the optimization mechanism (optimization of a cost function using a BVP) implies that the population modeled is a set of hyper-rational agents who have the same information regarding the final time horizon of the pandemic (e.g., when effective vaccines will be available) and relative costs of social distancing (e.g., how to value the cost of social distancing relative to the burden of the disease). While this framework is not meant to capture the complexity of heterogeneous human behaviors with bounded rationality nor the stochasticity of epidemics, it does provide insight into how rational people could behave *if* they had perfect foresight. In this setup, society’s time-varying social distancing strategy is not an input to the analysis but an output that emerges from the problem structure and model parameters. As we show later in the paper, we find that strategies that could be interpreted as suppression and mitigation strategies emerge from this simple model. There is a fast switch from mitigation to suppression strategies as the cost of social distancing and the time-horizon of the pandemic decreases.

## 2 Approach Overview

Here we provide a brief outline of our approach. Section 6 provides mathematical details that will be of interest to infectious disease modelers and mathematicians. We use a variational analysis approach, which is a mathematical technique that can be used to derive functions that minimize or maximize a quantity (such as a cost) over an interval, to combine a standard Susceptible-Infected-Recovered (SIR) epidemic model describing disease transmission with a cost function describing both costs of infections and costs of social distancing specified over the time horizon of interest. The SIR model is an evolutionary equation whereby the epidemiological state variables’ current values determine their rate of change. As long as the parameters that enter the SIR model remain constant, the present state variables determine the future evolution entirely. Using variational principles, model parameters can change dynamically to reach a pre-specified end state and time by selecting an epidemiological trajectory that minimizes the total cost of infections and social distancing. This approach converts the SIR model to a fixed time horizon model.

The state variables of the SIR model include the population densities of the susceptible (*s*), infected (*i*), and recovered (*r*) populations. The model parameters include a constant per capita disease progression rate *γ* from infected to recovered and the per capita disease transmission rate *β*. At the beginning of the epidemic without any behavioral adaptation, the transmission rate is constant *β*_0_, and the ratio *β*_0_/*γ* defines the basic reproduction number *R*_0_, which is an input parameter to our implementation of the model. In our model, the transmission rate *β*, and in turn the reproduction number, changes over time as determined by the variational approach. This change in transmission represents social distancing policies and behavioral changes. We define *R*_*D*_ to be the reproduction number that is modified by social distancing policy and resulting behavioral change. The variable *R*_*τ*_ is the effective reproduction number (*R*_*τ*_ = *sR*_*D*_) that captures both the change in the reproduction number due to behavioral changes *and* due to the removal of susceptibles from the population. We also define a cost parameter *c*, which describes the relative cost of social distancing compared to the cost of infection, and *τ* _final_, which describes the time at which the epidemic is expected to end. The time horizon can represent different types of prior expected durations, such as when an effective vaccine is made available or when the population is close to having reached herd immunity. Both *c* and *τ*_final_ are additional inputs to our model. The initial prevalence of the disease *i*_0_ is the final input, which determines the initial perceived risk of being infected, and together with *c* and *τ*_final_ determines the initial social mixing behaviors. Table 1 summarizes the input parameters to our model and provides the values we used in this study.

**Table 1:**
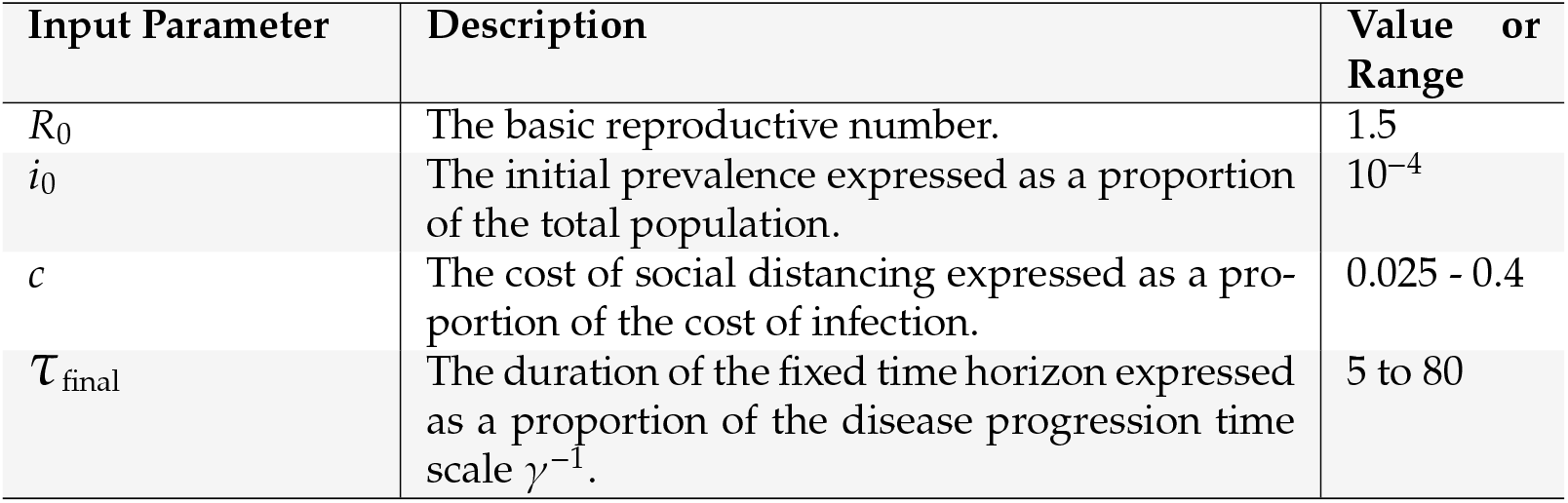
Input Parameters and their considered values.

### 2.1 Herd Immunity and Uncontrolled Epidemics

Interpretation of our final results is aided by understanding two conceptually important values of *s*. The first is *s*_∞_. This is the proportion of the population that would remain in a susceptible state at the end of an uncontrolled epidemic given that the epidemic started with a low initial infection rate (*i*_0_ ≪ 1) and that most individuals were susceptible to start (no individuals were in the recovered state or already immune to the disease at the start of the epidemic). The second important quantity is *s*_*H*_. This is the maximum proportion of individuals who could be in the susceptible state in a population that has achieved herd immunity. In other words, if *s*_*H*_ individuals are susceptible and the proportion of the population in the recovered state is 1 − *s*_*H*_, herd immunity will have been achieved. In other words, if a small number of infectious individuals were introduced into the population, the infection rate would decline rather than increase exponentially. In any case that *R*_0_ *>* 1, *s*_*H*_ *> s*_∞_. In other words, in an uncontrolled epidemic, fewer people will be in the susceptible state at the end (uninfected) than would be required to achieve herd immunity. The state variables of the model are summarized in table 2 together with those describing the time scales and herd-immunity.

**Table 2:** Model Variables.

| Variable | Description |
| --- | --- |
| $\tau$ | Dimensionless time (in units of $\gamma^{-1}$). |
| $i$ | Proportion of population in infectious state. |
| $s$ | Proportion of population in the susceptible state. |
| $s_H$ | Maximum possible proportion of population that remain in the susceptible state after reaching herd-immunity. |

## 3 Results

In this section, we first present results from our optimization analysis using the baseline parameter values. We then present the main outcomes of a wide range of optimization runs to characterize how the policy response is shaped by the time-horizon of the epidemic considered and the perceived cost of social distancing.

### 3.1 Mitigation and Suppression strategies emerge from different assumptions

In this paper, strategies are not an input to the analysis but an output and a result of the optimization procedure and the underlying assumptions. Instead of pre-specifying strategies to the model, we specify a theory-based mathematical description of how the disease progresses in the population (the SIR model), an objective function that includes the costs and benefits of social distancing, and let the optimization procedure determine the optimal time-varying distancing policy given relative costs and time-horizons of the pandemic. We then explore how different assumptions can lead to different strategies.

Figure 1 presents two different types of dynamics we observe as outcomes of our optimization model, which, for simplicity, we label as “mitigation” and “suppression” strategies. The left panel shows the disease dynamics of a mitigation strategy, and the right shows the dynamics of a suppression strategy. In the mitigation result, the levels of infection peak and then subside, and a substantial portion of the population eventually becomes infected. The mitigation strategy results from a set of parameters representing a relatively high cost of social distancing compared to infection cost. The suppression strategy results from a set of parameters representing a relatively low cost of social distancing compared to infection cost. Therefore, it is unsurprising that the mitigation strategy results in more infections than the suppression strategy. The final proportion of the population infected is less than what would occur if the epidemic spread with no mitigation (i.e., the number of susceptibles remains above the lower dashed red line). In addition, the total number of people who have become infected and have recovered is sufficient to produce herd immunity (i.e., the number of susceptibles is below the upper red line.) However, the suppression strategy does not allow the pandemic to take off and therefore only results in a modest number of infections towards the end of the time frame considered.

**Figure 1:**
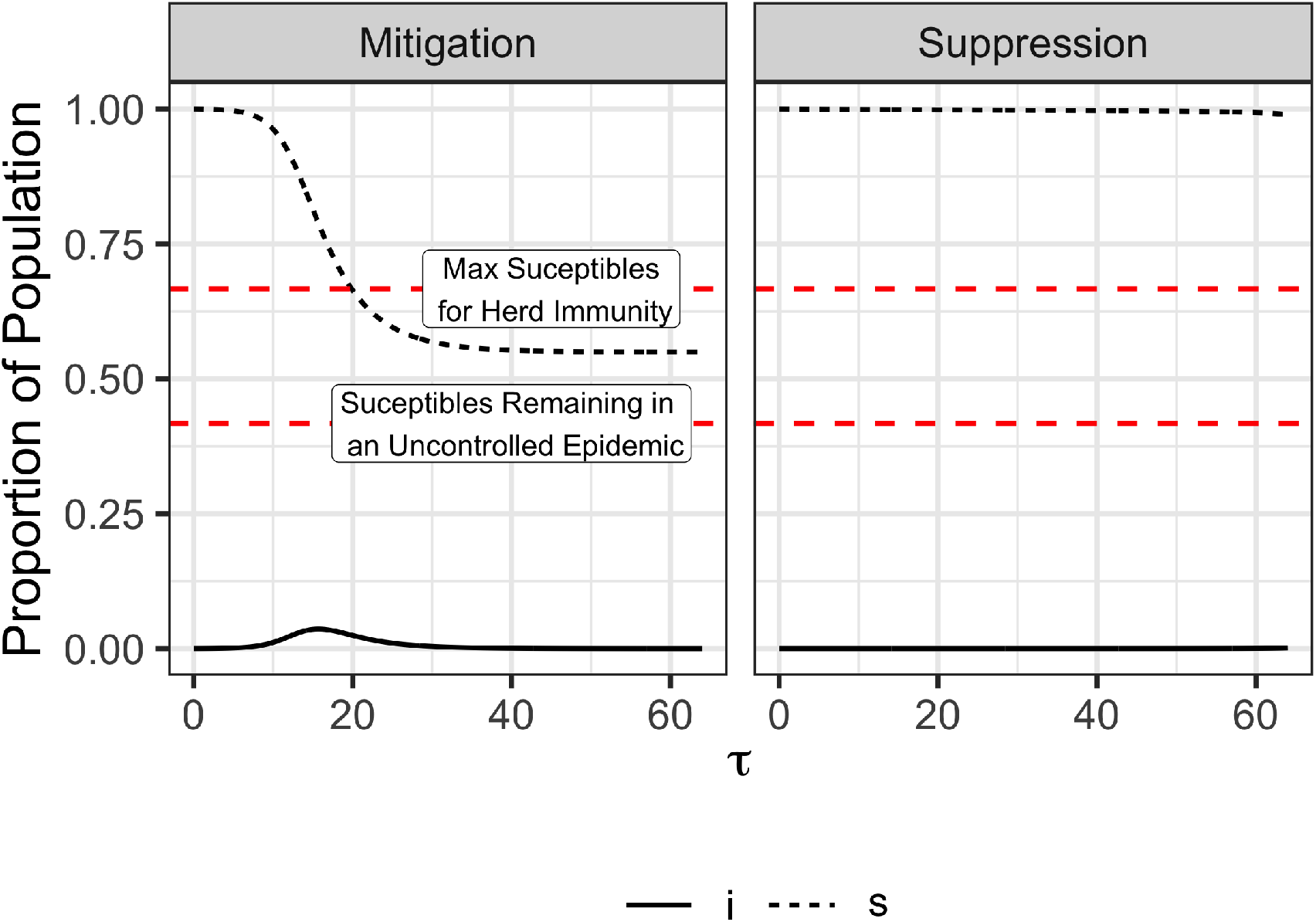
Mitigation vs. Suppression Disease Dynamics. In both the left and right panels, *R*_0_ = 1.5, *i*_0_ = 10^−4^, and τ_final_ = 64. The panels differ only in the value of *c*, the relative cost of social distancing compared to infections. The left panel shows the qualitative dynamics of a mitigation strategy. In this case *c* = 0.4. The right panel shows the qualitative dynamics of a suppression strategy. In this case *c* = 0.025. Recall that *R*_*D*_ is the social distancing reproduction number and *R*_τ_ is the reproduction number that accounts for decreases in the reproduction number due to both social distancing and removal of susceptibles from the population.

Figure 2 presents the same trajectories and policies discussed below but now demonstrate the dynamics of the time-varying reproductive number *R*_*D*_ and the effective reproductive number *R*_*τ*_ as a function of time for a case with resulting in a mitigation strategy and a case resulting in a suppression strategy. The mitigation strategy results in a dynamic reproduction number *R*_*D*_(τ) systematically above one, reflecting this policy’s unwillingness to control the epidemic, resulting from the relatively high value of *c*, the cost of social distancing compared to the cost of infection. Social distancing is the greatest (*R*_*D*_ is the lowest) approximately when infections peak (see Figure 1). In contrast, the suppression strategy keeps *R*_*D*_, and *R*_τ_ below one during the beginning of the pandemic but ultimately allows *R*_*D*_ *>* 1 once infection levels have become very low and once it is close enough to the end of the pandemic (*τ*_final_) that some growth in infections still results in very few infections throughout the pandemic. These results align closely with the observed behaviors of countries as vaccines have become increasingly available.

**Figure 2:**
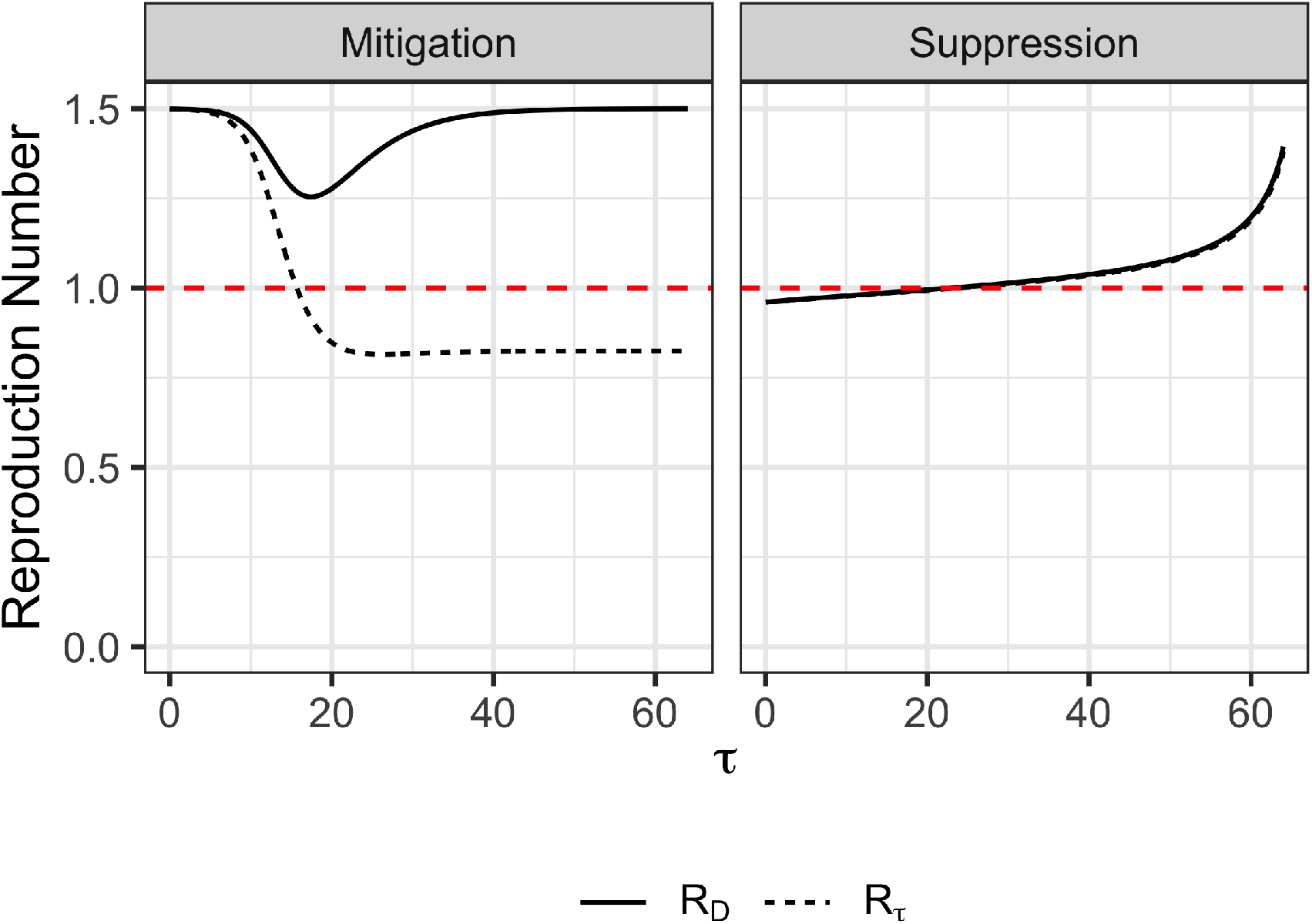
Mitigation vs. Suppression Strategies - *R*_*D*_ and *R*_*τ*_. In both the left and right panels, *R*_0_ = 1.5, *i*_0_ = 10^−4^, and τ_final_ = 64. The panels differ only in the value of *c*, the relative cost of social distancing compared to infections. The left panel shows the qualitative dynamics of a mitigation strategy; in this case *c* = 0.4. The right panel shows the qualitative dynamics of a suppression strategy; in this case *c* = 0.025.

Figure 3 shows *R*_*D*_ for a range of values of *τ*_final_ and *c*. For the values shown, when *τ*_final_ is less than or equal to 32, the dynamics of *R*_*D*_ are consistent with a suppression strategy. When *τ*_final_ increases to 64 and 128, solutions emerge that are consistent with a mitigation strategy, as shown in Figure 2. Figure 3 shows that the switch from mitigation to suppression strategy is fairly abrupt. For example, when *τ*_final_ = 64, the dynamics of *R*_*D*_ when *c* = 0.025 and 0.05 are suppression strategies and *c* = 0.2and0.4 are mitigation strategies. Only *c* = 0.1 appears to not strictly be either a mitigation or suppression strategy. In a “pure” mitigation strategy, herd immunity is achieved and full re-opening can occur (i.e., *R*_*D*_ = *R*_0_) before *τ*_final_. In this case, *R*_*D*_(*τ*_final_) *< R*_0_.

**Figure 3:**
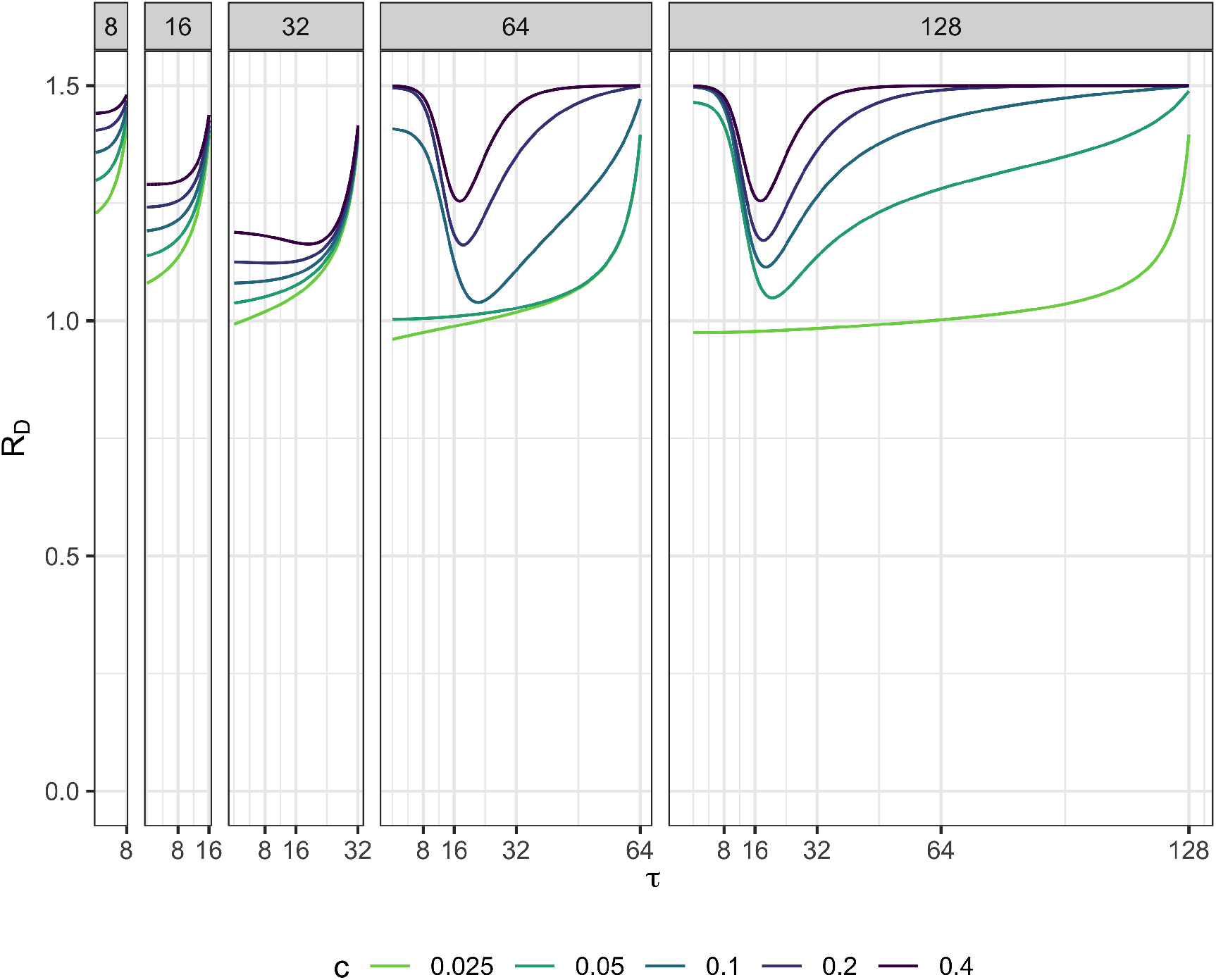
Optimal Distancing Policies. In all cases shown, *R*_0_ = 1.5 and *i*_0_ = 10^−4^. Panel names show the values of τ_final_ used in each analysis. Line colors show optimal social distancing policies for different values of *c*.

### 3.2 Optimal Strategy as a Function of Costs and Time-Frame

Figure 4 presents the epidemic size stemming from a range of policies when we vary the cost of social distancing *c* and the epidemic duration τfinal and perform 741 optimization runs. We use the epidemic size at the end of the epidemic time frame to indicate which policy was chosen. While we do not create or propose any artificial policy category scheme, the figure exhibits a clear discontinuity. The yellow region of the plot, with a long-time frame (e.g., vaccines take too long to develop) and high cost of social distancing (e.g., no income support is provided to families, and families must engage in infectious mixing to survive), result in a high epidemic size and thus reveal a mitigation strategy. Conversely, society chooses a suppression strategy with either a short epidemic time frame and/or low cost of social distancing. As figure 4 demonstrates, a deterministic SIR coupled with an analytical optimization framework can suggest very different strategies to manage an epidemic under alternative conditions.

**Figure 4:**
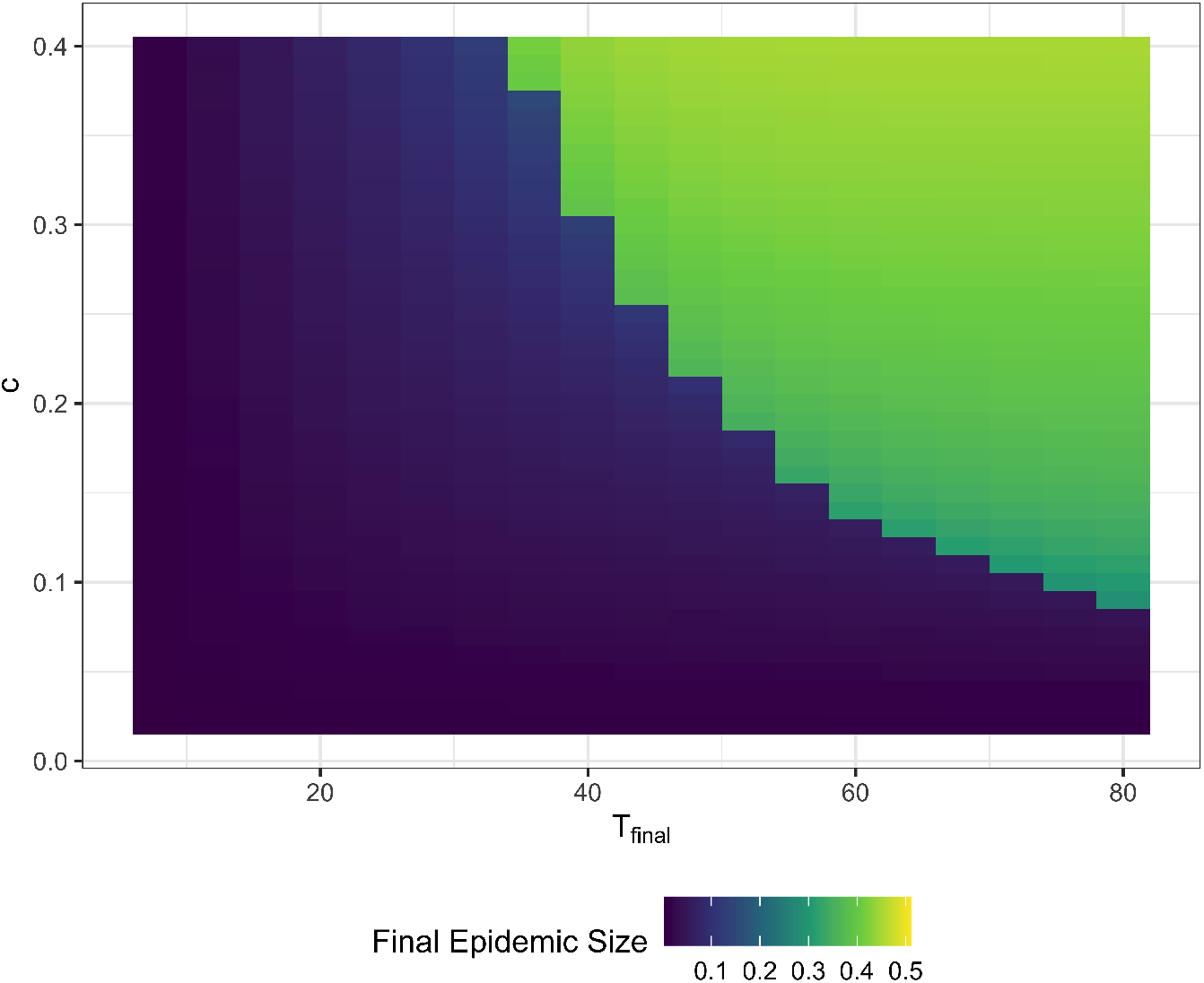
Final Epidemic Sizes for Optimal Policies. This plot shows the final epidemic sizes when optimal policies are followed for a range of values of time horizon, τ_final_ and relative cost of social distancing, *c*.

## 4 Discussion

The results presented previously revealed the characteristics of optimal social distancing policies under a variety of conditions by formulating the social distancing policy decision as a boundary value problem. In this set up, society decided to balance the costs and benefits of social distancing contingent on the costs of reducing transmission relative to the burden imposed by the disease. To the best of our knowledge, our paper is unique in using this approach. Because this approach does not impose a constraint in the functional form of the solution, social distancing strategies emerge from the problem structure and parameters rather than being imposed *a priori*. The next paragraphs discuss the main findings from these computational experiments. We find that the relative time-horizon of the pandemic (i.e., the time it takes to develop effective vaccines and treatments) and the relative cost of social distancing influence the optimal policy choice. We also find that optimal suppression and mitigation strategies have characteristics that may seem counter-intuitive and can challenge common wisdom.

### Perceived time-horizons and relative costs shape society’s response to the pandemic

The most salient finding from these experiments was that the perceived time-horizon and costs of social distancing provoked a fast switch from suppression to a mitigation strategy. The higher the costs of social distancing and the longer the epidemic time horizon, the higher the chance society will pursue a mitigation strategy prioritizing livelihoods over saving lives. This sharp, non-linear switch from mitigation to suppression might be a challenging aspect to address in a pandemic with characteristics that fall close to the edge of the frontier revealed in figure 4. Because countries with different socioeconomic characteristics and populations with heterogeneous values may perceive or experience social distancing costs differently, any pandemic that falls close to that frontier will inevitably result in disagreement and lack of coordination.

### Optimal Strategies were smooth and decisive in an idealized setting

In this mathematical exercise, society’s response to the pandemic was smooth and decisive, meaning that policymakers did not start with blunt lockdowns only to reverse course a few weeks later and to instate restrictions again. Therefore, our model does not reproduce results in policies seen in the real world, including adaptive policies that imposed and lifted social distancing orders adaptively, which were tested in our previous work [17] or strategies that use fast periodic switching between suppression and mitigation [18, 19]. One can interpret this result in two ways. One possibility is that our analytical machinery cannot reproduce these policies, and they are indeed better than the smooth, optimal policies we obtained. Alternatively, the smooth policies revealed in this study are indeed optimal, and never have been tried. This is nuanced and may be a stark finding. First, suppose decisive strategies have never been tried. In that case, there is no chance that empirical analysis will reveal the true difference between current strategies and ideal strategies because an appropriate counterfactual is not available in the data. Second, modeling studies that simulated a limited set of heuristic policies might present a menu of dominated options. This aspect should be investigated with further analysis, specifically with more nuanced models that include heterogeneity and stochasticity. Suppose such smooth policies are, in fact optimal even under more challenging conditions. In that case, investigating the performance of these policies in more realistic models could prove a useful and worthwhile exercise.

### Optimal suppression strategies do not hold R_*τ*_ *<* 1 indefinitely, and optimal mitigation strategies do not allow completely uncontrolled epidemics

Early in the COVID-19 pandemic, the common-wisdom view was that society ought to keep COVID-19 cases from growing, and economists have gone a step further to propose that society ought to maximize utility, keeping *R*_*τ*_ *<* 1 [20]. While this seems a reasonable statement, our findings do not corroborate the idea that *R*_*τ*_ should be below one *indefinitely at all costs*. As noted in figures 1 and 2, *R*_*τ*_ eventually surpasses one before the end of the epidemic time-frame under a suppression strategy, even under the scenarios that were most favorable to a suppression strategy. These results, therefore, suggest that *R*_*τ*_ *<* 1 is not a permanent end-goal of optimal suppression strategies. Instead, optimal suppression strategies balance the costs and benefits of social distancing measures by keeping *R*_*τ*_ *<* 1 at the beginning of the epidemic and only allow a controlled increase in infections when they are the least dangerous - towards the end of the epidemic. Nevertheless, an indefinite zero-covid strategy was not found to be desirable from a welfare maximization standpoint using this simple model. Instead, the optimal strategy was contingent and time-varying - it did imply **near** zero-infections at the beginning of the epidemic, and the latter led to a modest and controlled rise in infections. This finding aligns with how New Zealand and other countries that successfully adopted aggressive strategies to curb COVID-19 have proceeded. Countries that adopted zero-covid strategies at the beginning of the COVID-19 pandemic later transitioned to relaxing their policies once vaccines were widely available. This finding also is in line with other studies which found that robust COVID-19 reopening strategies had time-varying reopening thresholds with more stringent thresholds at the start of the pandemic and less stringent thresholds towards the end of the pandemic [17, 21].

Our findings also revealed that no optimal mitigation strategies imply a completely uncontrolled epidemic. This finding suggests that static transmissibility was never found to be an optimal strategy. This result demonstrates that static transmissibility *β*_*τ*_ is never an optimal policy. While this is a characteristic of the traditional SIR model, these results demonstrate that a rational society will always seek to minimize the worst costs of pandemics through *some* distancing, especially when mitigation measures are inexpensive. This finding blurs the lines between mitigation and suppression strategies in the sense that no optimal mitigation and suppression strategies found in this exercise is the *extreme* version of those policies. Optimal “mitigation” strategies involved some distancing measures, and optimal “suppression” strategies always involved some level of infection.

### Poor communication and misinformation will push society towards mass numbers of infections and deaths in a new pandemic

Assuming full rationality from people is a strong, demanding assumption. However, we interpret and offer our results in a positive rather than a normative tone. Instead of asserting which strategy society should take, we claim that *if agents behaved rationally, then under idealized conditions, this is how they would behave*. Under this positive interpretation, we find that *even a rational society will allow massive infections* if they believe that the costs of social distancing are too high relative to the cost of infection, or that the pandemic and vaccines will take too long to develop or may never be developed. We also find that infections are allowed to rise towards the end of the pandemic even when a suppression strategy is followed. This simple mathematical model, coupled with an optimization routine, demonstrates that it is critical to properly communicate the timelines involved with mitigation strategies and the relative costs of infection and social distancing.

### Uncertainty, complexity, and heterogeneity make it difficult to use this framework to support decision-making in the next pandemic

Based on our findings, one might imagine that a simplified optimization framework such as the one used in this paper will provide a rational approach to support decision-making during the next pandemic. If the costs of social distancing relative to the costs of infection are known and if the time-horizon to a vaccine and the disease parameters are set, and if the model structure is correct, then this framework could provide rational decision support to provide optimal policies. However, we warn the reader that none of these assumptions will hold during the onset of the next pandemic. While the optimization structure used in this paper allows for mathematical tractability, both the costs and the time-horizon of a pandemic are uncertain, heterogeneous quantities. Pandemics will be relatively more costly to societies that cannot mitigate the economic costs of social distancing with income support policies financed by debt. Similarly, wealthy nations will have access to treatments and vaccines before other nations. The result is that different countries will be positioned at different regions in the parameter space we presented. This heterogeneity predictably could cause countries to be forced into different strategies. Moreover, within-country dynamics can also play an important role, with different jurisdictions adopting different strategies. The corollary of these factors is that the world *will not* adopt a single best social distancing strategy, and chaotic behavior should be expected. In light of these findings, however, one can still resort to decision-making approaches that seek to find policies that are robust to these uncertain factors but achieve acceptable performance [17].

## 5 Conclusions

The COVID-19 pandemic has affected billions of people worldwide and has been unprecedented in scale and duration. During the early stages of the pandemic and in the absence of a vaccine, policymakers had to take extraordinary measures, implementing a range of nonpharmaceutical public health interventions (NPIs) to mitigate deaths caused by the spread of the highly transmissible virus (SARS-CoV-2). These measures included mask-wearing and social distancing policies that ranged from partial closings of business to complete lockdown.

Despite the deluge of research on COVID-19 policy, the effectiveness of policies as applied to COVID-19 will only partially inform the decision to suppress or mitigate the next pandemic. A new pandemic may bring surprises, different epidemiological characteristics, and the evidence created for COVID-19 might not translate directly to new pathogens with different properties. The intuitive and potentially misleading inclination is to learn what worked for COVID-19 and immediately apply those lessons to a new pandemic. An alternative and complementary learning mode are to understand the fundamental properties of the decision problem we faced in the COVID-19 pandemic and ask how epidemiological and economic parameters should determine our choices in a new pandemic.

As we have seen in the COVID-19 pandemic, social distancing remains a critical intervention to mitigate the spread and deaths caused by a novel, highly transmissible infectious disease. However, prolonged social distancing can lead to significant declines in social well-being and widespread economic hardships and uncertainties. Under these circumstances, policymakers need to make the discomforting decision of social distancing intervention policies that make a tradeoff between lives and livelihoods.

In this paper, we investigated how the choice of the strategy to govern Nonpharmaceutical interventions change depending on society’s assessment of the time-horizon of the pandemic (i.e., the time it takes to obtain and distribute an effective vaccine) and the costs of suppression measures relative to the social cost of infections. Even assuming that society was uniform and governed by a rational, hyper-rational agent, we have found that society could choose a suppression strategy if it believes that social distancing is too costly or if the pandemic would take too long to curb with vaccines or highly effective treatments. These results point to the importance of public messaging. They could provide an internally consistent mathematical explanation of why nonpharmaceutical interventions have enjoyed low popularity and adherence in some areas of the world.

## 6 Methods

### 6.1 The SIR Model

We use a standard Susceptible, Infectious, Recovered (SIR) model of disease transmission. The standard SIR model describes disease transmission in a well-mixed system in which an interaction between any two individuals is equally likely. While the solutions and non-dimensional form of the standard SIR model have been extensively studied, we show the derivation of the dimensionless SIR model for clarity as we will later show both the dimensional and dimensionless forms of cost functions and solutions to the social distancing problems.

The differential equations describing the dynamics of the susceptible, infectious, and recovered individuals in the population are:

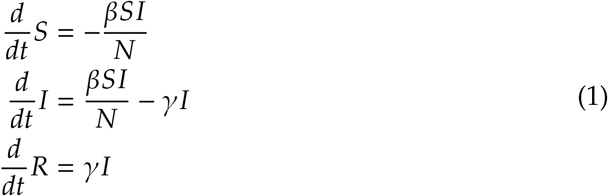

where *S* is the number of the susceptible, *I* is the number of the infectious, *R* is the number of the recovered, and *N* is the total number of individuals in the population. The parameters *β* and *γ* respectively represent the transmission and recovery rate and have units of inverse time.

#### 6.1.1 Non-Dimensionalization

As is typically done, we define the dimensionless unit of time *τ*, which is:

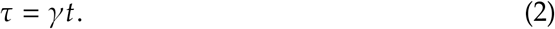

In other words, in the dimensionless SIR model, *τ* = 1 represents the average duration an individual is infectious for. We further recast the state variables in their dimensionless form where

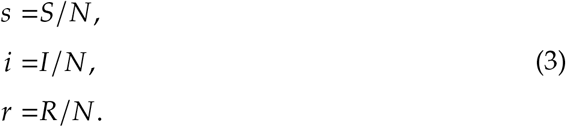

The dimensionless differential equations for the population dynamics become:

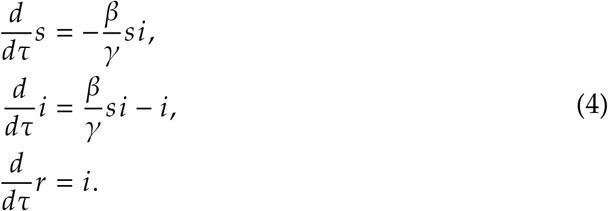

By defining the reproduction number 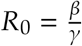, these equations can be formulated as

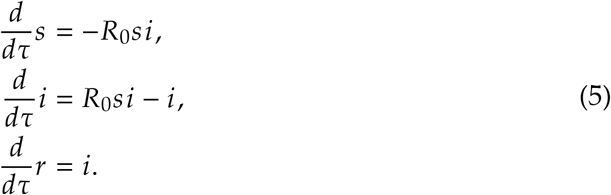

#### 6.1.2 Final Epidemic Size

Without mitigation measures, and under the assumption that the initial conditions for the proportion of the population infected is very small (i.e., *i*(*τ* = 0) ≪ 1), and nearly the entire population is initially susceptible (i.e., *s*(*τ* = 0) ≈ 1), the proportion of the population that remains susceptible as *τ* → ∞ is given by the solution to the equation [22]

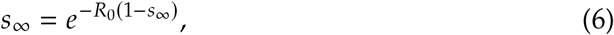

and the final epidemic size is 1 − *s*_∞_.

#### 6.1.3 The Effective Reproduction Number and Herd Immunity

The effective reproduction number *R*_*τ*_ describes the average number of infections produced by each infected individual in the population at time *τ*. In the standard SIR model without mitigation measures,

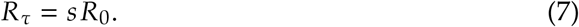

Herd immunity occurs when the number of those susceptible in the population is reduced either by natural infection or vaccination such that the effective reproduction number is less than 1. Therefore, the maximum proportion of susceptible individuals in a population with herd immunity is

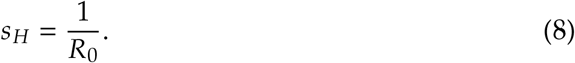

### 6.2 Time-Dependent Transmission

In classic SIR models, *β* and *R*_0_ are constant parameters. This paper considers the case where the transmission rate, *β*, is dynamic and influenced by behavioral changes. We will use the notation that *β* is a dynamic variable and *β*_0_ is the transmission rate without any behavioral mitigation. Similarly, we define *R*_*D*_(*τ*) as the dynamic reproduction number, which is a function of social distancing behavior. *R*_0_ is the reproduction number in the absence of any behavioral mitigation. In a dynamic framework, the dimensionless SIR model, Equations 5 become:

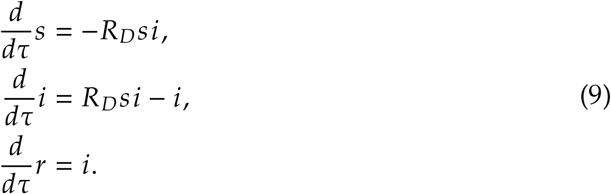

In the framework with a dynamic transmission rate, the effective reproduction number becomes:

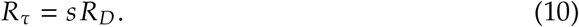

### 6.3 Cost Functions

We want to find the social distancing policy that minimizes a total cost, which includes the cost of social distancing and the cost of infections over a pre-specified time horizon ranging from time *t* = 0 to *t* = *t*_final_. The social distancing policy is described by *β*(*t*), the transmission rate, that depends on social distancing behavior at time *t*. We further define *β*_0_ to be the infection rate for the disease in the absence of any behavioral change. Therefore, *β* (*t*) is restricted to the interval (0, *β*_0_] The total cost is:

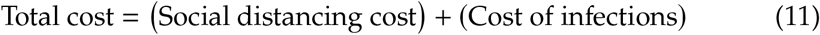

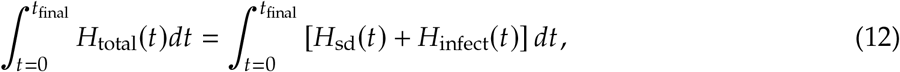

where *H*_sd_(*t*) is the cost of social distancing per unit time, and *H*_infect_ is the cost of infections. As the transmission rate *β*(*t*) decreases, the cost of social distancing per unit time, *H*_sd_(*t*), increases, but the cost of infections per unit time *H*_infect_(*t*) decreases.

#### 6.3.1 Cost of Infections

The number of new infections per unit time is 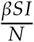, which comes from the SIR equations. If we define *D* to be the average cost of infection per infected individual. Then, the cost per unit time of infections is:

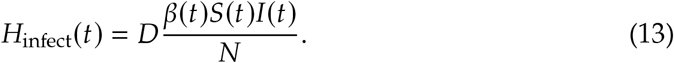

Note that the total cost of infections is:

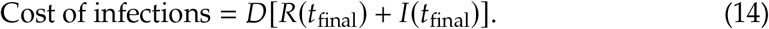

#### 6.3.2 Cost of social distancing

We assume that the total cost of social distancing is proportional to the size of the population *N*, and a cost parameter *C*, which has units cost per person per unit time. Therefore, we define the cost per unit time of social distancing to be:

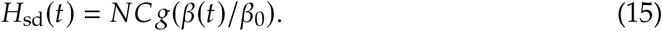

Here, *g*(*x*) is the function that defines the relationship between the relative cost of social distancing and relative reduction in the transmission parameter. The function *g*(*x*) should have the following properties:

1. **g**(**x**) **is a monotonically decreasing function on the interval** [0, 1]. The theoretical setup of the problem determines this property. The cost of social distancing should increase as the transmission rate is further decreased.
2. lim_**x**→**0**_+ **g**(**x**) = ∞. In theoretical terms, it is not possible to stop all transmission completely; therefore, the cost of decreasing transmission rates to zero should be infinite. From a practical perspective, this restriction will prevent optimal solutions to *β*(*t*) from passing through *β* (*t*) = 0, and if the initial condition for *β* (*t*) is positive, the solution will remain positive.
3. 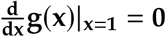. Theoretically, if there is no cost of infections, the optimal *β* (*t*) = *β*_0_. Therefore, the cost of social distancing should be minimized when 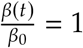, From a practical perspective, combined with condition 2, this condition ensures that solutions to *β* (*t*) will be bounded to the interval (0, *β*_0_] if correctly initialized to the interval.
4. **g(1)=0**. When 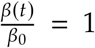 there are no behavioral changes so the cost of social distancing should be zero.

Based on these three required properties, we choose the function to have the form

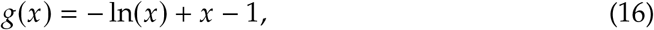

and thus, substituting this into equation 15, the cost per unit time of social distancing is

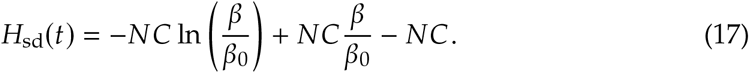

#### 6.3.3 Dimensionless Cost Equations

We now derive the dimensionless form of the cost equation. We define a dimensionless cost of social distancing, *c* to be:

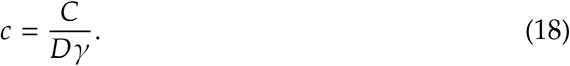

Effectively, *c* compares the cost of social distancing to the cost of infection. Recall that *C* has dimensions of cost per individual per unit time, *D* has dimensions of cost per individual, and *γ* has units of inverse time. Therefore, *c* is dimensionless. When *c* is small, people will more willingly social-distance as they feel that the cost of becoming ill outweighs the benefits of mixing socially. Their preference favors their lives and health compared to livelihoods and non-health-related well-being. When *c* is large, people’s preferences are switched. We define the dimensionless cost:

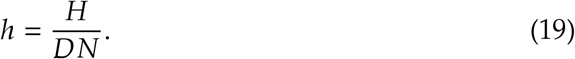

We can make the following substitutions in all equations:

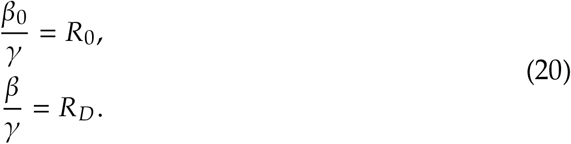

Then, the dimensionless cost of infection, Equation 13, becomes:

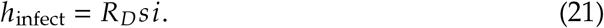

The dimensionless cost of social distancing, Equation 15, becomes:

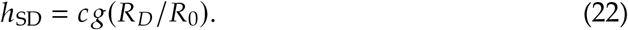

We denote the cost function as *h*(*s, i, R*_*D*_) which can be expressed as:

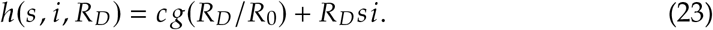

By setting *τ*_final_ = *γ τ*_final_, the total cost in its dimensionless form is

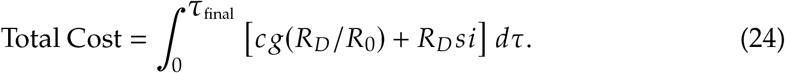

### 6.4 Full Optimization

Calculus of variations is used to find the functions *y*_*i*_(*τ*) that maximize or minimize

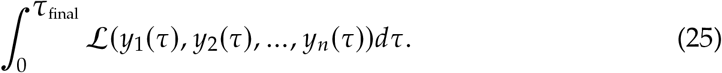

The functions *y*_*i*_ [23] that satisfy the following conditions are extrema of the integral in Equation 25. The conditions are given by:

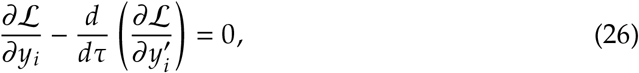

where 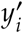 indicated the derivative 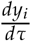.

Our main objective is to solve for *R*_*D*_ that minimizes Equation 23. Because the integrand is a function not only of *R*_*D*_, but of *s* and *i* as well, we introduce the functional Lagrange multipliers *λ*_*i*_ and *λ*_*s*_ to ensure that the conditions of the differential equations governing the infection dynamics (Equations 5) are met. Then, based on the constraints provided by the SIR dynamics, the full Lagrangian becomes:

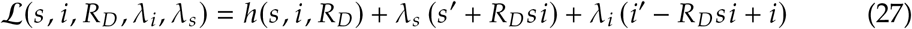

Note that we do not need to include a differential equation constraint for *r* since the equations for *s* and *i* ensure that *r* = 1 − *s* − *i*.

We can now apply Equation 26 to all *y*_*i*_ ∈ {*s, i, R*_*D*_, *λ*_*i*_, *λ*_*s*_}.

*y*_*i*_ = *s* yields:

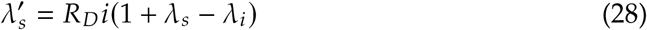

*y*_*i*_ = *i* yields:

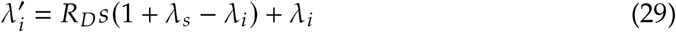

*y*_*i*_ = *R*_*D*_ yields:

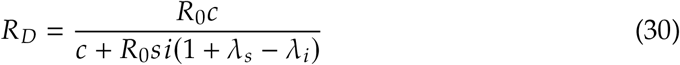

Note that applying *y*_*i*_ = *λ*_*s*_ and *y*_*i*_ = *λ*_*i*_ to Equation 26 returns the constraint differential equations for *s* and *i* from Equation 5.

The differential equations can then be solved as a two-point boundary value problem. For all cases presented in this paper, we set *i*(0) = *i*_0_, *s*(0) = 1 − *i*_0_, *λ*_*s*_(*τ*_final_) = 0, and *λ*_*i*_(*τ*_final_) = 0. We set *λ*_*s*_ and *λ*_*i*_ to be equal to zero at the end point of the interval because *s*(*τ*_final_) and *i*(*τ*_final_) are unconstrained. We solved the 2-point boundary value problems in R using the bvpSolve package.[24]

## Data Availability

This is a computational study and does not contain emperical data.

## Contributions

- Sarah Nowak (Ph.D.) led the effort in conceptualizing and formulating the model structure and led the overall modeling effort, including model implementation. Sarah is the corresponding author of this study and can be contacted at:
- Pedro Nascimento de Lima (MSc) contributed to the model analysis and interpretation and drafting and revising the manuscript.
- Raffaele Vardavas (Ph.D.) contributed to the model formulation, implementation, and interpretation of model outputs and drafting and revising the manuscript.

## Acknowledgements

We wish to thank the National Cancer Institute (R21CA157571), and the National Institute of Allergies and Infectious Diseases (R01AI118705 & R01AI160240) for providing support in projects that led to preliminary work and ideas that motivated this project. Dr. Nowak acknowledges support from the Blodwen S. Huber Early Career Green and Gold Professor in Pathology and Laboratory Medicine at The Robert Larner, M.D. College of Medicine.

## Disclaimer

The results and conclusions drawn on this working paper have not been peer-reviewed and do not necessarily represent the opinions of the RAND Corporation.

